# Early Prediction of Post-TAVR Left Ventricular Remodeling Using CT-Derived Radiomics and Clinical Variables

**DOI:** 10.64898/2026.05.28.26354361

**Authors:** Mostafa Rezaeitaleshmahalleh, Shahab Masoumi, Fatemeh Razaviamri, Amir Rouhollahi, Edoardo Zancanaro, Tommaso Hinna Danesi, Brian C. Ayers, Arminder Jassar, Ashraf Sabe, Farhad R. Nezami

## Abstract

**Background:** Adverse left ventricular (LV) remodeling after transcatheter aortic valve replacement (TAVR) is associated with impaired functional recovery and adverse long-term outcomes, yet imaging-based risk stratification remains limited.

**Objectives:** This study sought to determine whether CT-derived radiomic and geometric myocardial features, integrated with procedural and clinical variables, can predict adverse LV remodeling after TAVR.

**Methods:** We retrospectively analyzed 232 consecutive TAVR recipients with paired pre- and post-procedural LV mass index (LVMI) measurements. Adverse remodeling was defined as a ≥10% increase in LVMI at follow-up. Pre-procedural CT was used to derive three-dimensional LV geometric descriptors, ray-tracing wall-thickness metrics, and myocardial texture radiomic features. Random forest classifiers were developed across six models of sequentially increasing complexity.

**Results:** Adverse LV remodeling occurred in 52 patients (22.4%). Geometry-only model showed limited discrimination (AUC 0.62), whereas wall-thickness radiomics substantially improved performance (AUC 0.84). A multimodal pre-procedural model combining CT radiomics with pre-procedural LVMI, residual valve insufficiency, and prior coronary revascularization achieved an AUC of 0.86 (95% CI 0.73 to 0.98). Addition of post-procedural mean transvalvular gradient further improved discrimination (AUC 0.91, 95% CI 0.81 to 0.98). SHAP analysis identified post-procedural mean aortic gradient and radiomic markers of myocardial heterogeneity as the leading predictors.

**Conclusions:** CT-derived radiomic characterization of myocardial heterogeneity provides incremental prognostic information beyond conventional geometric assessment for identifying patients at risk of adverse LV remodeling after TAVR. These findings extend the role of pre-procedural CT beyond anatomical planning toward quantitative myocardial phenotyping and individualized risk stratification, although prospective validation is required to establish clinical utility.

## 1. Introduction

Aortic stenosis (AS) is the most prevalent valvular heart disease in industrialized countries, with rising prevalence as populations age^1,2^. Chronic pressure overload drives compensatory left ventricular (LV) hypertrophy that ultimately becomes maladaptive, progressing to fibrosis, diastolic and systolic dysfunction, heart failure, and increased mortality^3,4^.

Transcatheter aortic valve replacement (TAVR) has revolutionized the management of severe AS by providing a less invasive alternative to surgery. Relief of valvular obstruction after TAVR frequently yields favorable reverse remodeling, with regression of LV mass, reduced wall thickness, and improved diastolic function^5,6^. However, post-TAVR remodeling is heterogeneous. Many patients exhibit incomplete or maladaptive reverse remodeling characterized by persistent hypertrophy, ventricular dilatation or progressive fibrosis, which is associated with reduced functional recovery, higher rates of heart-failure hospitalization, and worse long-term survival^7,8^.

Early identification of patients at risk for maladaptive remodeling may improve risk stratification and post-procedural management. Current prediction strategies rely largely on demographic, clinical, and global functional metrics such as LV ejection fraction or global longitudinal strain^9–11^. Although clinically useful, these measures incompletely characterize the spatial and microstructural heterogeneity of myocardial remodeling ^12^.

Pre-procedural cardiac computed tomography (CCT), routinely acquired for TAVR planning, provides detailed information beyond conventional anatomical assessment. In addition to defining LV geometry, CCT enables quantification of regional hypertrophy and myocardial textural heterogeneity that may reflect underlying structural remodeling. Emerging evidence suggests that radiomic and geometric imaging biomarkers may improve prognostic assessment^13^. however, prior studies have largely evaluated these features independently, and an integrated framework combining geometry, texture, wall-thickness heterogeneity, and clinical variables remains lacking.

We therefore hypothesized that combining pre-procedural myocardial geometric descriptors, radiomic texture features, and relevant clinical variables would enable accurate ML-based prediction of post-TAVR LV remodeling trajectories. To test this hypothesis, we reconstructed patient-specific LV models from pre-procedural CT scans, extracted three-dimensional geometric and radiomic metrics, and integrated these imaging-derived biomarkers with clinical data to train and evaluate ML models. This approach aimed to establish an imaging-informed, quantitative framework for risk assessment that leverages routinely acquired CT data to improve patient selection, support clinical decision-making, and inform personalized post-TAVR follow-up.

## 2. Materials and Methods

### 2.1. Study Population and Data Acquisition

A total of 933 contrast-enhanced pre-TAVR CTA studies performed at Brigham and Women’s Hospital were retrospectively identified under Institutional Review Board approval (2010P000292). All procedures complied with institutional guidelines and the Declaration of Helsinki. Patients with available paired LVMI measurements obtained before TAVR and at 12–14 months post-procedure were included to enable assessment of LV remodeling. After applying inclusion criteria, 232 patients comprised the final study cohort.

Patients were stratified according to post-TAVR LVMI change into: 1) an adverse remodeling group with a ≥10% increase in LVMI and 2) a stable/minimal remodeling group with a *<*10% LVMI change, irrespective of direction. This threshold is supported by prior TAVR-specific outcome studies, including the PARTNER trials, demonstrating that failure to achieve LV mass regression at 1 year is independently associated with a 71% higher risk of death or rehospitalization over 5-year follow-up, and that persistent severe hypertrophy post-TAVR carries significant prognostic weight.^7,14,15^

### 2.2. Geometric Characterization of LV Morphology and Myocardial Architecture

LV myocardial segmentation was obtained using Cardiovison^16^, an in-house AI-based platform, with details in our prior publication and the Supplement(Section. S2). These segmentations were used for subsequent geometric, wall-thickness, and radiomic analyses.

#### 2.2.1. Morphometric Analysis of the Endocardial Surface

Geometric descriptors were extracted from the segmented endocardial surface. Fundamental metrics included enclosed cavity volume (*V*), surface-to-volume ratio, and maximum and minimum diameters (*D*_max_, *D*_min_). Regional shape was further characterized by additional morphometric indices^17^.

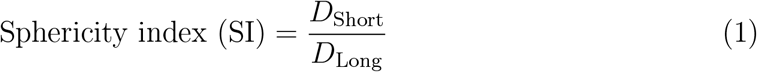

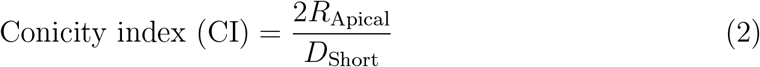

As shown in Figure 1a, the LV long axis was defined as the distance from the midpoint of the mitral valve plane to the apex (LV height); the short-axis diameter (*D*_Short_) was measured perpendicular to the long axis at its midpoint. Conicity was the ratio of apical to short-axis diameter, with the apical radius (*R*_Apical_) taken as the radius of the best-fitting sphere at the apex. Geometric irregularity was quantified by the undulation index (UI), non-sphericity index (NSI), and ellipticity index (EI)^18^.

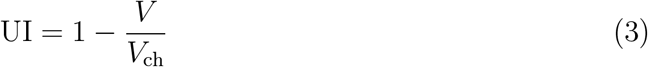

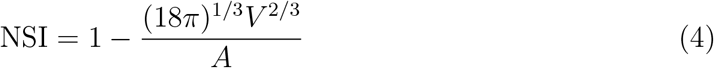

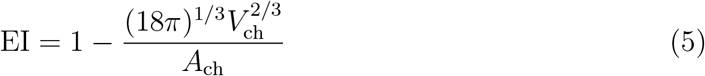

*V*_ch_ and *A*_ch_ denote the volume and surface area of the convex hull enclosing the LV (Figure 1b), a minimal convex reference used to quantify surface protrusions, indentations, and overall shape complexity. Together, these metrics characterized both global LV configuration and regional geometric distortion.

**Figure 1:**
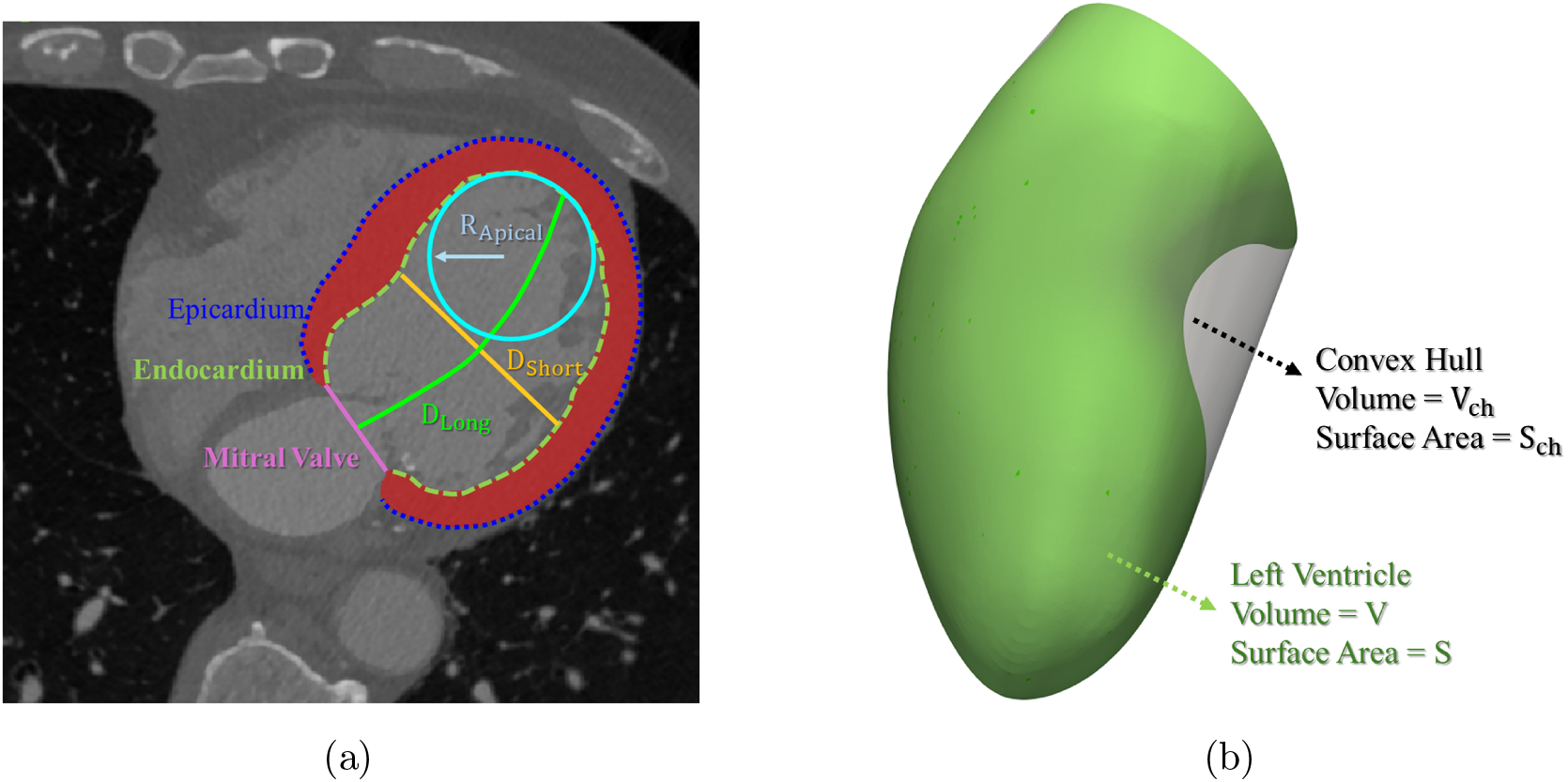
Geometric characterization of the left ventricle from cardiac CT. (a) Cross-sectional CT image showing the extracted left ventricular geometric parameters. (b) Three-dimensional left ventricular model illustrating ventricular and convex hull volume and surface area measurements.

#### 2.2.2. Ray-Tracing–Based Quantification of Myocardial Wall Thickness

Wall thickness was quantified by ray-tracing on the three-dimensional myocardial geometry reconstructed in Section 2.2. The ventricular centerline was extracted and uniformly resampled with the Vascular Modeling Toolkit (VMTK). At each centerline point, a local orthonormal frame was constructed: the tangent vector was estimated from adjacent points, and two perpendicular basis vectors (*u, v*) were defined relative to a consistent global reference. Within the (*u, v*) plane, radial rays were cast at angular increments from 0 to 2*π* on a 100 *×* 100 grid (100 longitudinal *×* 100 circumferential) (Figure 2a).

**Figure 2:**
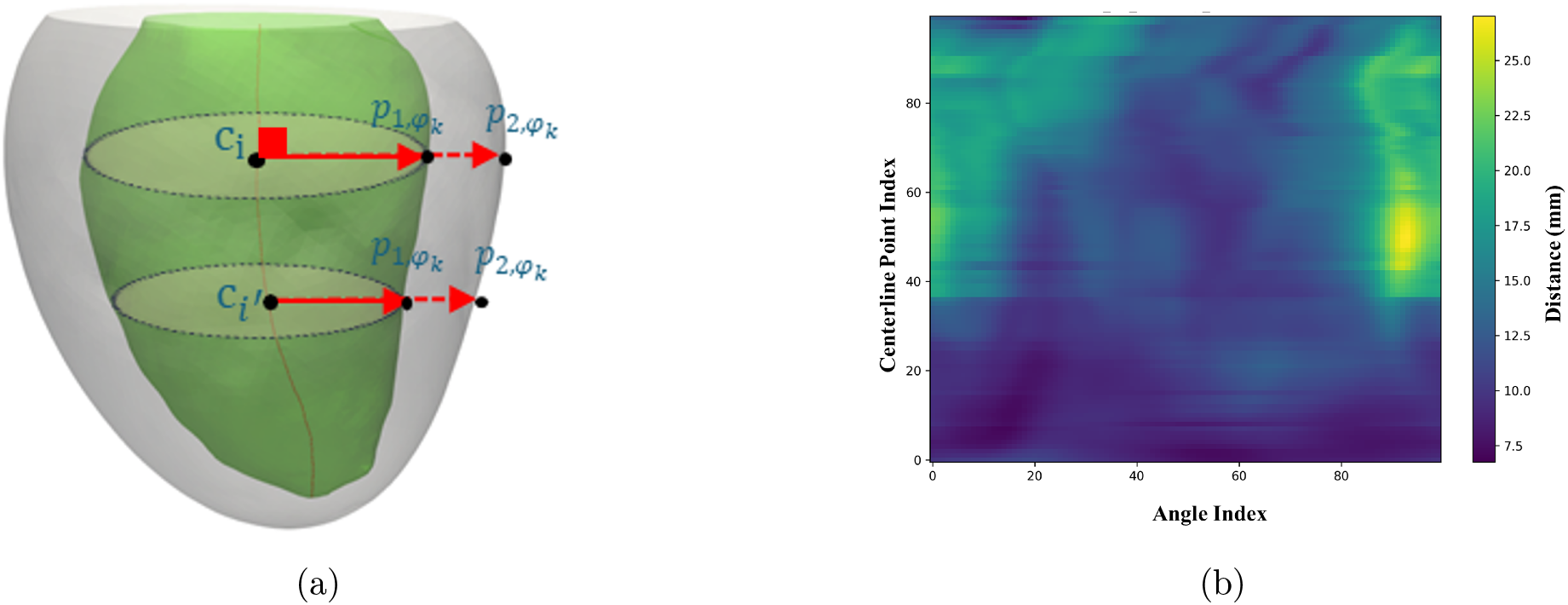
Workflow for ray-tracing–based myocardial wall-thickness quantification. (A)Construction of local coordinate frames and radial ray casting around each centerline point. (B)Generation of a 2D wall-thickness map parameterized by centerline position and circumferential angle.

Along each ray, the first and second intersections with the myocardial mesh were taken as the endocardial and epicardial surfaces, and local wall thickness was the Euclidean distance between them. Measurements were assembled into a 2D wall-thickness map parameterized by longitudinal position and circumferential angle (Figure 2b). Missing values (*<*10%) were imputed by nearest-neighbor interpolation to preserve spatial continuity.

To characterize spatial heterogeneity in myocardial wall-thickness distribution, radiomic features were extracted from the 2D thickness maps using PyRadiomics v3.1.0^19^ with `force2D=True`, yielding first-order statistics and higher-order texture features from the gray-level co-occurrence matrix (GLCM), gray-level run length matrix (GLRLM), gray-level size zone matrix (GLSZM), gray-level dependence matrix (GLDM), and neighboring gray tone difference matrix (NGTDM).

### 2.3. Radiomics-Based Textural Analysis of the Myocardium

To characterize tissue-level heterogeneity within the myocardium, radiomic features were extracted directly from the 3D CTA intensity distribution using the segmented myocardium geometry as the region of interest.

Both the myocardial mask and the corresponding CTA volume were resampled to an isotropic voxel size of 0.2 *×* 0.2 *×* 0.2 mm^3^ using B-spline interpolation implemented in SimpleITK. Using the same PyRadiomics framework, features were extracted from the 3-dimensional myocardial volume, including first-order intensity statistics and second-order texture features from GLCM, GLRLM, GLSZM, GLDM, and NGTDM. Additional details regarding preprocessing, feature definitions, and extraction parameters have been described previously.^20^

### 2.4. Predictive Modeling and Interpretability

Following univariate dimension reduction and feature selection (Supplementary material Section. S3), a random forest classifier was chosen as the primary model because of its robustness and established utility in clinical prediction tasks. Hyperparameters were optimized by grid search using AUC as the primary criterion. The dataset was partitioned into stratified training (80%) and held-out testing (20%) cohorts to preserve outcome prevalence. Within the training cohort, 5-fold stratified cross-validation was used for model tuning and selection. Model performance was evaluated exclusively on the held-out test set, which was not used during model training or feature selection, to reduce the risk of overfitting. Predictive performance was assessed primarily by AUC, with additional reporting of accuracy, sensitivity, specificity, and F1-score. Model interpretability was evaluated using SHapley Additive exPlanations (SHAP),^21,22^ which quantify the relative contribution of each feature to model output.

## 3. Results

### 3.1. Patient Characteristics

Adverse LV remodeling occurred in 52 patients (22.4%), while 180 (77.6%) demonstrated favorable remodeling at follow-up. Baseline demographic and clinical characteristics were well balanced between groups (Table 1). Sex distribution, prevalence of prior PCI, CABG, myocardial infarction, and aortic valve annular calcification were comparable between cohorts (all *p* > 0.10). LV ejection fraction ≥50% was preserved in the majority of patients in both groups (86.1% vs. 75.9%, *p* = 0.087), and aortic valve morphology was predominantly tricuspid (90.6% vs. 92.3%, *p* = 0.880). Pre-procedural hemodynamic indices of stenosis severity, including mean aortic gradient, peak aortic valve velocity, and effective valve area, did not differ significantly between groups, confirming comparable baseline stenosis burden. Pre-procedural LVMI was significantly higher in the favourable remodeling group (116.85 ± 30.41 vs. 95.39 ± 23.88 g/m^2^; *p* < 0.0001), indicating that patients who subsequently developed adverse remodeling carried a lower baseline hypertrophic burden. Following the procedure, effective valve area and peak aortic valve velocity were equivalent between groups; however, post-procedural mean aortic gradient was significantly higher in the favourable remodeling group (12.67 ± 5.56 vs. 10.41 ± 4.04 mmHg; *p* = 0.011).

**Table 1:**
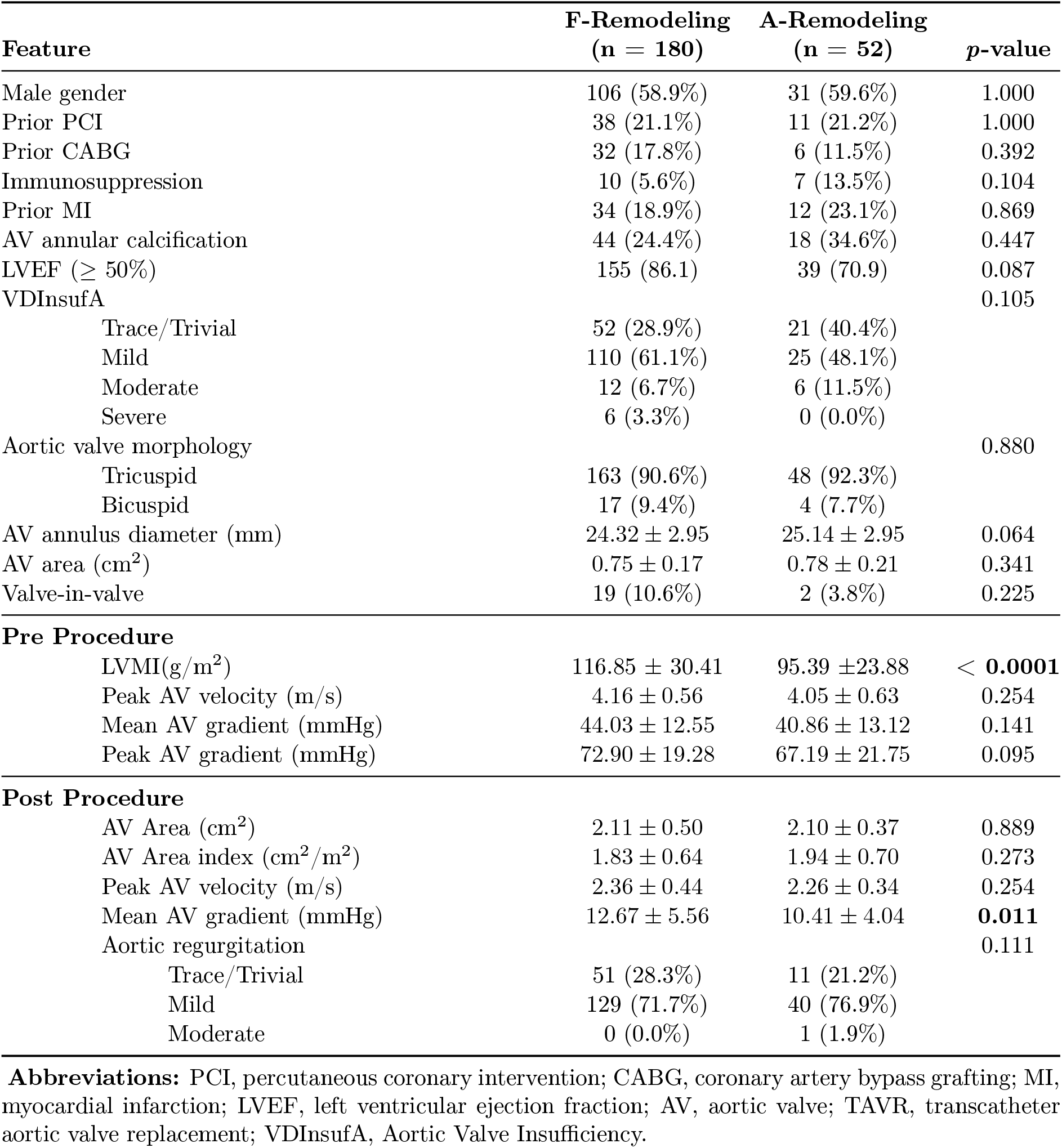
Baseline demographic, clinical, and echocardiographic characteristics of the study cohort. Study cohort stratified by Favorable Remodeling (F-Remodeling) group and Adverse Remodeling (A-Remodeling) group. Continuous variables are reported as mean ± SD and categorical variables as *n* (%).

| Feature | F-Remodeling<br>(n = 180) | A-Remodeling<br>(n = 52) | p-value |
| --- | --- | --- | --- |
| Male gender | 106 (58.9%) | 31 (59.6%) | 1.000 |
| Prior PCI | 38 (21.1%) | 11 (21.2%) | 1.000 |
| Prior CABG | 32 (17.8%) | 6 (11.5%) | 0.392 |
| Immunosuppression | 10 (5.6%) | 7 (13.5%) | 0.104 |
| Prior MI | 34 (18.9%) | 12 (23.1%) | 0.869 |
| AV annular calcification | 44 (24.4%) | 18 (34.6%) | 0.447 |
| LVEF ($\geq 50\%$) | 155 (86.1) | 39 (70.9) | 0.087 |
| VDInsufA |  |  | 0.105 |
| Trace/Trivial | 52 (28.9%) | 21 (40.4%) |  |
| Mild | 110 (61.1%) | 25 (48.1%) |  |
| Moderate | 12 (6.7%) | 6 (11.5%) |  |
| Severe | 6 (3.3%) | 0 (0.0%) |  |
| Aortic valve morphology |  |  | 0.880 |
| Tricuspid | 163 (90.6%) | 48 (92.3%) |  |
| Bicuspid | 17 (9.4%) | 4 (7.7%) |  |
| AV annulus diameter (mm) | 24.32 ± 2.95 | 25.14 ± 2.95 | 0.064 |
| AV area (cm<sup>2</sup>) | 0.75 ± 0.17 | 0.78 ± 0.21 | 0.341 |
| Valve-in-valve | 19 (10.6%) | 2 (3.8%) | 0.225 |
| **Pre Procedure** |  |  |  |
| LVMI(g/m<sup>2</sup>) | 116.85 ± 30.41 | 95.39 ± 23.88 | < **0.0001** |
| Peak AV velocity (m/s) | 4.16 ± 0.56 | 4.05 ± 0.63 | 0.254 |
| Mean AV gradient (mmHg) | 44.03 ± 12.55 | 40.86 ± 13.12 | 0.141 |
| Peak AV gradient (mmHg) | 72.90 ± 19.28 | 67.19 ± 21.75 | 0.095 |
| **Post Procedure** |  |  |  |
| AV Area (cm<sup>2</sup>) | 2.11 ± 0.50 | 2.10 ± 0.37 | 0.889 |
| AV Area index (cm<sup>2</sup>/m<sup>2</sup>) | 1.83 ± 0.64 | 1.94 ± 0.70 | 0.273 |
| Peak AV velocity (m/s) | 2.36 ± 0.44 | 2.26 ± 0.34 | 0.254 |
| Mean AV gradient (mmHg) | 12.67 ± 5.56 | 10.41 ± 4.04 | **0.011** |
| Aortic regurgitation |  |  | 0.111 |
| Trace/Trivial | 51 (28.3%) | 11 (21.2%) |  |
| Mild | 129 (71.7%) | 40 (76.9%) |  |
| Moderate | 0 (0.0%) | 1 (1.9%) |  |

**Abbreviations:** PCI, percutaneous coronary intervention; CABG, coronary artery bypass grafting; MI, myocardial infarction; LVEF, left ventricular ejection fraction; AV, aortic valve; TAVR, transcatheter aortic valve replacement; VDInsufA, Aortic Valve Insufficiency.

### 3.2. LV Geometric Indices

Global LV size metrics such as chamber volume and surface-to-volume ratio were not significantly different between groups (*p >* 0.6). The minimal endocardial radius, however, was larger in the adverse-remodeling group (7.4 ± 2.4 mm vs. 6.2 ± 2.4 mm; *p <* 0.001), indicating loss of the narrowest ventricular curvature. Patients with adverse remodeling also exhibited more globular and conical chambers, as reflected by higher sphericity (0.78 ± 0.13 vs. 0.75 ± 0.18; *p* = 0.038) and conicity indices (0.64 ± 0.16 vs. 0.54 ± 0.15; *p* < 0.0001). In contrast, undulation, non-sphericity, and ellipticity indices were not significantly different(Table 2). Taken together, these findings suggest that regional geometric remodeling, rather than global cavity size, differentiates adverse from favorable post-TAVR adaptation.

**Table 2:** Comparative measurement of LV geometrical features. Study cohort stratified by Favorable Remodeling (F-Remodeling) group and Adverse Remodeling (A-Remodeling) group Parameters list as mean ± SD for each group and variables with p-value*<*0.05 are in bold.

| Feature | F-Remodeling | A-Remodeling | *p-value* |
| --- | --- | --- | --- |
| Volume (mm<sup>3</sup>) | 1.20E5 ± 4.85E4 | 1.23E5 ± 5.07E4 | 0.672 |
| surface-to-volume ($\frac{1}{mm}$) | 0.122 ± 0.079 | 0.115 ± 0.021 | 0.711 |
| Minimum Radius (mm) | 6.185 ± 2.352 | 7.426 ± 2.405 | **<0.001** |
| Maximum Radius (mm) | 21.737 ± 3.529 | 22.092 ± 3.776 | 0.300 |
| UI | 0.064 ± 0.090 | 0.051 ± 0.028 | 0.569 |
| NSI | 0.304 ± 0.057 | 0.299 ± 0.022 | 0.846 |
| EI | 0.281 ± 0.020 | 0.282 ± 0.020 | 0.841 |
| SR | 0.752 ± 0.184 | 0.778 ± 0.129 | **0.038** |
| CI | 0.543 ± 0.146 | 0.641 ± 0.159 | **<0.0001** |

**UI:** undulation index, **NSI:** non-sphericity index, **EI:** Ellipticity Index, **CI:** Conicity Index and **SR:** Sphericity Index

### 3.3. Myocardial Wall-Thickness Radiomics

Analysis of ray-tracing–derived wall-thickness maps demonstrated that global summary statistics such as range, mean, median, and variance of thickness did not differ significantly between groups, implying similar overall thickness distributions. In contrast, discriminatory information resided in higher-order texture descriptors emphasizing regions of thin myocardium. Low Gray Level Run Emphasis, Small Area Low Gray Level Emphasis, and related Gray Level Dependence Matrix indices were higher in the adverse-remodeling group (all *p <* 0.05), reflecting a greater prevalence of spatially contiguous or clustered regions of thin wall (Table 3). These findings suggest that adverse remodeling is characterised by patchy regional thinning rather than diffuse myocardial atrophy.

**Table 3:** Radiomic features derived from ray-tracing–based 2D myocardial wall-thickness maps. Study cohort stratified by Favorable Remodeling (F-Remodeling) group and Adverse Remodeling (A-Remodeling) group. Parameters are reported as mean ±SD for each group, and variables with *p<* 0.05 are shown in bold.

| Feature | F-Remodeling | A-Remodeling | *p*-value |
| --- | --- | --- | --- |
| **First-order** |  |  |  |
| Range | 18.848 ± 5.397 | 17.549 ± 5.250 | 0.104 |
| 90Percentile | 15.673 ± 2.156 | 15.239 ± 1.901 | 0.154 |
| Maximum | 21.812 ± 4.089 | 21.074 ± 3.601 | 0.231 |
| TotalEnergy | 1.48E6 ± 3.67E5 | 1.40E6 ± 2.93E5 | 0.242 |
| RootMeanSquared | 12.103 ± 1.484 | 11.803 ± 1.263 | 0.242 |
| Mean | 11.739 ± 1.540 | 11.478 ± 1.217 | 0.253 |
| RobustMeanAbsoluteDeviation | 1.520 ± 0.610 | 1.407 ± 0.390 | 0.279 |
| InterquartileRange | 3.548 ± 1.397 | 3.305 ± 0.979 | 0.313 |
| **GLCM** |  |  |  |
| Autocorrelation | 109.139 ± 61.643 | 93.268 ± 56.886 | 0.126 |
| Idn | 0.989 ± 0.003 | 0.989 ± 0.003 | 0.128 |
| JointAverage | 9.531 ± 3.161 | 8.753 ± 3.047 | 0.143 |
| ClusterShade | 96.615 ± 204.356 | 55.459 ± 165.753 | 0.435 |
| **GLRLM** |  |  |  |
| LowGrayLevelRunEmphasis | 0.025 ± 0.022 | 0.029 ± 0.023 | **0.043** |
| LongRunLowGrayLevelEmphasis | 3.661 ± 13.644 | 3.079 ± 3.550 | 0.082 |
| HighGrayLevelRunEmphasis | 128.038 ± 69.451 | 109.172 ± 64.001 | 0.089 |
| RunVariance | 24.618 ± 14.801 | 25.499 ± 9.555 | 0.156 |
| ShortRunHighGrayLevelEmphasis | 41.777 ± 30.884 | 35.639 ± 28.480 | 0.194 |
| **GLSZM** |  |  |  |
| SmallAreaLowGrayLevelEmphasis | 0.004 ± 0.003 | 0.005 ± 0.003 | **0.024** |
| LargeAreaEmphasis | 48.321 ± 21.014 | 49.911 ± 17.453 | 0.375 |
| LargeAreaHighGrayLevelEmphasis | 4168.045 ± 2717.348 | 3723.938 ± 2510.334 | 0.389 |
| **NGTDM** |  |  |  |
| Strength | 1.126 ± 0.935 | 0.946 ± 0.772 | 0.134 |
| Busyness | 1.527 ± 1.413 | 1.827 ± 1.623 | 0.160 |
| Complexity | 33.561 ± 22.179 | 28.751 ± 18.563 | 0.160 |
| Contrast | 0.009 ± 0.004 | 0.009 ± 0.004 | 0.237 |
| **GLDM** |  |  |  |
| SDLGLE | 0.006 ± 0.008 | 0.007 ± 0.005 | **0.034** |
| LDLGLE | 0.297 ± 0.504 | 0.307 ± 0.248 | **0.045** |
| LowGrayLevelEmphasis | 0.039 ± 0.060 | 0.040 ± 0.032 | **0.046** |
| SDHGLE | 24.550 ± 15.570 | 20.767 ± 14.507 | 0.102 |
| HighGrayLevelEmphasis | 109.546 ± 61.783 | 93.696 ± 57.071 | 0.126 |
| LDHGLE | 714.403 ± 400.616 | 614.943 ± 372.241 | 0.146 |

SmallDependenceLowGrayLevelEmphasis: SDLGLE, LargeDependenceLowGrayLevelEmphasis: LDLGLE, SmallDependenceHighGrayLevelEmphasis: SDHGLE, LargeDependenceHighGrayLevelEmphasis: LDHGLE

### 3.4. CT Texture Radiomics

Radiomic features extracted from the three-dimensional CT volume demonstrated increased myocardial heterogeneity in patients with adverse remodeling. First-order features revealed lower minimum Hounsfield unit values and higher kurtosis (*p <* 0.05), indicating a broader and more heterogeneous distribution of attenuation values with more extreme low and high intensities. GLCM features, including Sum Average, Joint Average, Difference Variance, and Cluster Prominence were significantly elevate (*p <* 0.05), consistent with more pronounced clustering of disparate tissue densities. GLRLM and GLSZM features emphasising high gray-level runs and zones (e.g., Short Run High Gray Level Emphasis, Long Run High Gray Level Emphasis and Small Area High Gray Level Emphasis) were also significantly elevated (all *p <* 0.05), while Gray Level Dependence measures showed increased dependence on high-density voxels with reduced non-uniformity (Table 4). Collectively, these findings are consistent with increased spatial heterogeneity and clustering of high-attenuation regions, potentially reflecting focal fibrosis or calcification within the myocardium of patients who develop adverse remodeling.

**Table 4:** Texture radiomic features extracted from the CT image data within the defined volume of interest. Study cohort stratified by Favorable Remodeling (F-Remodeling) group and Adverse Remodeling (A-Remodeling) group. Parameters list as mean± SD for each group and variables with p-value*<*0.05 are in bold.

| Feature | F-Remodeling | A-Remodeling | *p-value* |
| --- | --- | --- | --- |
| **Firstorder** |  |  |  |
| Variance | 2629.451 ± 1563.919 | 3070.190 ± 2304.860 | 0.112 |
| Range | 1404.911 ± 654.705 | 1502.346 ± 853.825 | 0.380 |
| Minimum | -307.472 ± 180.696 | -385.769 ± 333.085 | **0.027** |
| Kurtosis | 25.308 ± 45.440 | 43.456 ± 82.351 | **0.040** |
| Energy | 5.15E10 ± 3.14E10 | 4.60E10 ± 2.77E10 | 0.251 |
| 10Percentile | 48.472 ± 22.955 | 45.385 ± 22.556 | 0.392 |
| **GLCM** |  |  |  |
| SumSquares | 4.014 ± 2.389 | 4.608 ± 3.312 | 0.151 |
| SumAverage | 34.785 ± 14.365 | 40.711 ± 26.286 | **0.035** |
| JointAverage | 17.392 ± 7.182 | 20.356 ± 13.143 | **0.035** |
| DifferenceVariance | 0.868 ± 0.428 | 1.098 ± 1.133 | **0.027** |
| Contrast | 1.743 ± 0.955 | 1.955 ± 1.383 | 0.207 |
| ClusterTendency | 14.312 ± 8.930 | 16.478 ± 12.161 | 0.159 |
| ClusterProminence | 11009.427 ± 43599.269 | 76246.186 ± 264171.665 | **0.002** |
| **GLRLM** |  |  |  |
| SRHGLE | 257.517 ± 266.792 | 398.269 ± 649.656 | **0.021** |
| RunLengthNonUniformity | 1143453.048 ± 519105.837 | 1018480.704 ± 420102.722 | 0.113 |
| LRHGLE | 1489.754 ± 2040.650 | 2915.001 ± 7747.595 | **0.027** |
| HGLRE | 360.184 ± 375.557 | 591.291 ± 1094.241 | **0.017** |
| GrayLevelVariance | 5.136 ± 2.854 | 5.975 ± 4.165 | 0.096 |
| GrayLevelNonUniformity | 369123.854 ± 148049.138 | 331978.519 ± 122447.243 | 0.100 |
| **GLSZM** |  |  |  |
| ZoneVariance | 6.56E7 ± 7.70E7 | 5.16E7 ± 4.72E7 | 0.215 |
| SAHGLE | 176.270 ± 190.806 | 267.913 ± 392.596 | **0.021** |
| SmallAreaEmphasis | 0.379 ± 0.057 | 0.391 ± 0.056 | 0.186 |
| SZNUN | 0.152 ± 0.038 | 0.160 ± 0.039 | 0.180 |
| LAHGLE | 1.82E10 ± 2.98E10 | 4.53E10 ± 1.81E10 | 0.056 |
| LargeAreaEmphasis | 6.57E7 ± 7.71E7 | 5.16E7 ± 4.72E7 | 0.215 |
| HGLZE | 415.777 ± 406.232 | 658.908 ± 1124.245 | **0.016** |
| GrayLevelVariance | 29.668 ± 22.372 | 41.763 ± 58.729 | **0.025** |
| GrayLevelNonUniformity | 4531.100 ± 2416.069 | 4131.632 ± 2179.865 | 0.285 |
| **NGTDM** |  |  |  |
| Complexity | 1246.638 ± 1534.712 | 1450.682 ± 1970.007 | 0.431 |
| Coarseness | 0.000 ± 0.000 | 0.000 ± 0.000 | 0.167 |
| Busyness | 237.574 ± 201.512 | 206.760 ± 193.564 | 0.328 |
| **GLDM** |  |  |  |
| SDHGLE | 11.191 ± 16.386 | 14.154 ± 17.692 | 0.260 |
| LDHGLE | 48650.551 ± 66151.089 | 97013.611 ± 250292.218 | **0.020** |
| HighGrayLevelEmphasis | 358.165 ± 375.158 | 588.700 ± 1092.866 | **0.018** |
| GrayLevelVariance | 4.287 ± 2.502 | 4.989 ± 3.673 | 0.114 |
| GrayLevelNonUniformity | 674386.090 ± 314583.535 | 604111.173 ± 246871.995 | 0.139 |
| DependenceNonUniformity | 247945.735 ± 94450.665 | 219355.406 ± 77536.162 | **0.047** |

SRHGLE: ShortRunHighGrayLevelEmphasis,SAHGLE: SmallAreaHighGrayLevelEmphasis,LRHGLE: LongRunHighGrayLevelEmphasis,LAHGLE: LargeAreaHighGrayLevelEmphasis,SZNUN: SizeZoneNonUniformityNormalized, SDHGLE:SmallDependenceHighGrayLevelEmphasis,HGLZE: HighGrayLevelZoneEmphasis, LDHGLE: LargeDependenceHighGrayLevelEmphasis, HGLRE: HighGrayLevelRunEmphasis,

### 3.5. Prediction Model Performance

Six models of increasing complexity, integrating LV geometric descriptors, CT radiomics, and procedural and clinical variables, were evaluated (Tables 5 and 6). ROC curves for all six are shown in Figure 3. The geometry-only model (Model 1) demonstrated limited discriminative performance (AUC 0.62 [95% CI: 0.36–0.87]), despite high specificity (0.97), indicating that geometric descriptors alone are not sufficient to capture remodeling risk. Myocardial texture radiomics alone (Model 2) improved discrimination (AUC 0.74 [95% CI: 0.54–0.90]), whereas a thickness-only radiomic model (Model 3) yielded the highest performance amongst single-domain models (AUC 0.84 [95% CI: 0.70–0.95]), albeit at the cost of reduced sensitivity (0.20). Combining complementary radiomic domains such as texture and thickness (Model 4) achieved an accuracy of 85% with highest specificity (1.00) and an AUC of 0.83 [95% CI: 0.67–0.96], supporting the additive discriminative value of multimodal radiomic characterization.

**Table 5:**
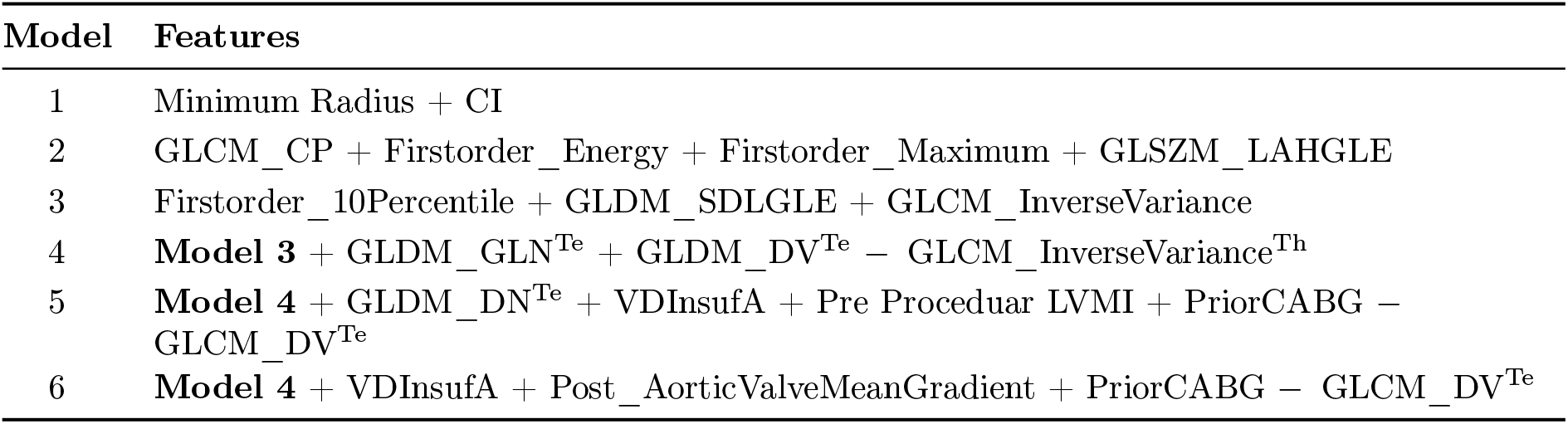
Overview of the predictive models and feature combinations evaluated in this study. Model (1) includes LV geometry, Model (2) myocardial texture, Model (3) myocardial thickness, and Model (4) combined texture-thickness radiomics. Models (5) and (6) include feature sets without and with post-procedural clinical/procedural variables, respectively. In Models 4–6, superscripts ^Te^ and ^Th^ denote radiomic features derived from myocardial texture and myocardial thickness, respectively.

**Table 6:** Classification performance of predictive models based on LV geometry, myocardial thickness, texture features, and PHI. The highest value in each column is highlighted in **bold**.

| Model# | Accuracy | F1-score | Sensitivity | Specificity | AUC |
| --- | --- | --- | --- | --- | --- |
| 1 | 0.85 | 0.53 | 0.40 | 0.97 | 0.62[0.36-0.87] |
| 2 | 0.81 | 0.47 | 0.40 | 0.95 | 0.74[0.54-0.90] |
| 3 | 0.81 | 0.31 | 0.20 | 0.97 | 0.84[0.70-0.95] |
| 4 | 0.85 | 0.46 | 0.30 | **1.00** | 0.83[0.67-0.96] |
| 5 | **0.89** | **0.67** | **0.50** | **1.00** | 0.86[0.73-0.98] |
| 6 | 0.85 | 0.59 | **0.50** | 0.95 | **0.91[0.81-0.98]** |

**Figure 3:**
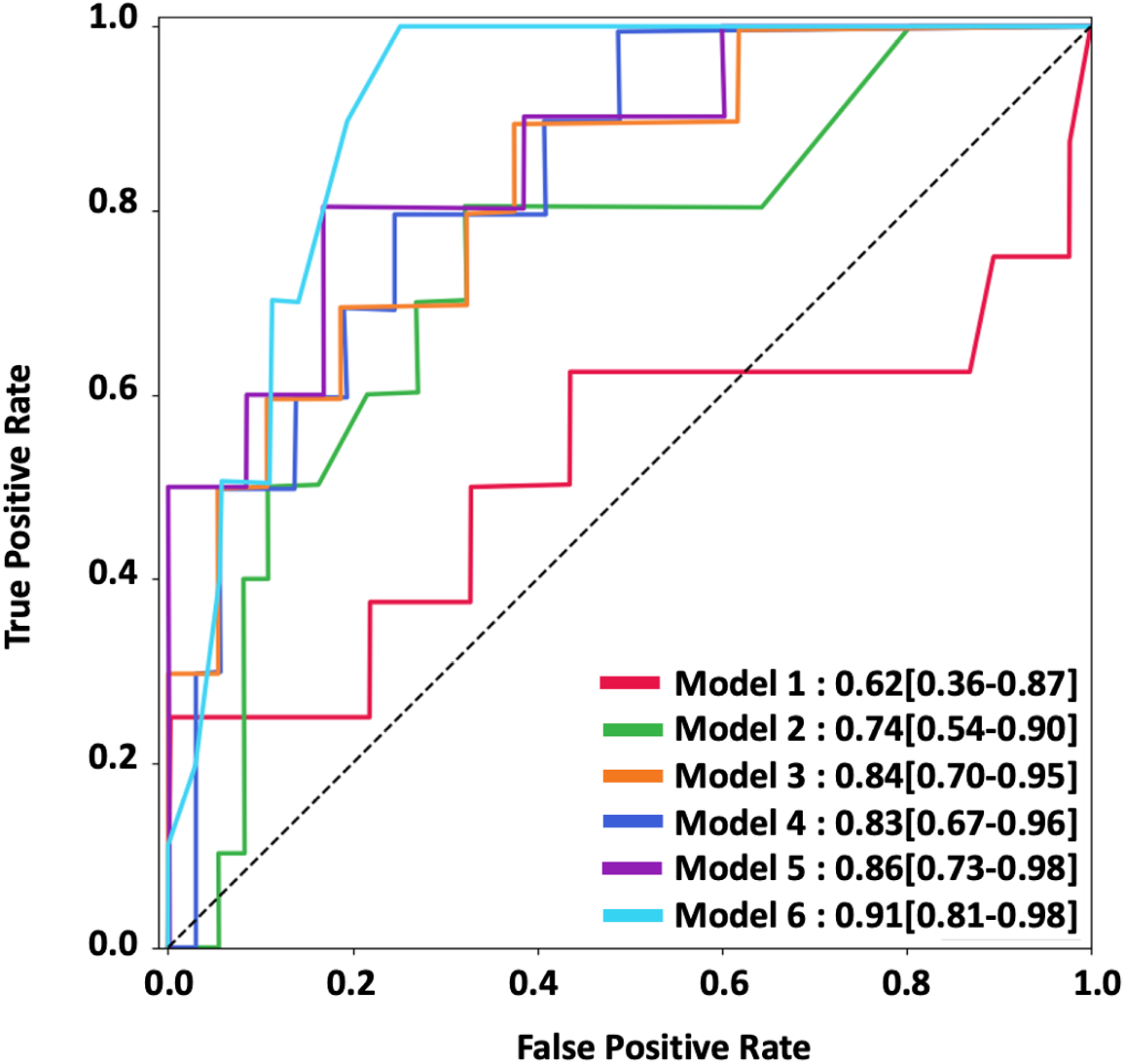
ROC Curves for the Six Predictive Models.

Augmentation with pre-procedural clinical variables including prior CABG history, residual valve insufficiency, and pre-procedural LVMI (Model 5) further improved performance (AUC 0.86 [95% CI: 0.73–0.98]; accuracy 89%; F1-score 0.67; sensitivity 0.50; specificity 1.00), demonstrating the complementary contribution of imaging and clinical features. The highest overall discrimination was achieved by Model 6, which incorporated post-procedural mean aortic gradient (AUC **0.91** [95% CI: 0.81–0.98]; accuracy 85%; F1-score 0.59; sensitivity 0.50; specificity 0.95), underscoring the dominant prognostic contribution of residual transvalvular hemodynamic burden to adverse LV remodeling following TAVR.

### 3.6. Feature Importance and Model Interpretability

SHapley Additive exPlanations (SHAP) analysis was used to interpret the final model (Figure 4). Post-TAVR mean transvalvular gradient emerged as the strongest predictor, with higher values associated with a greater likelihood of favorable remodeling. Radiomic features reflecting wall-thickness heterogeneity and myocardial texture variation were the next most influential predictors, followed by lower-percentile wall-thickness metrics.

**Figure 4:**
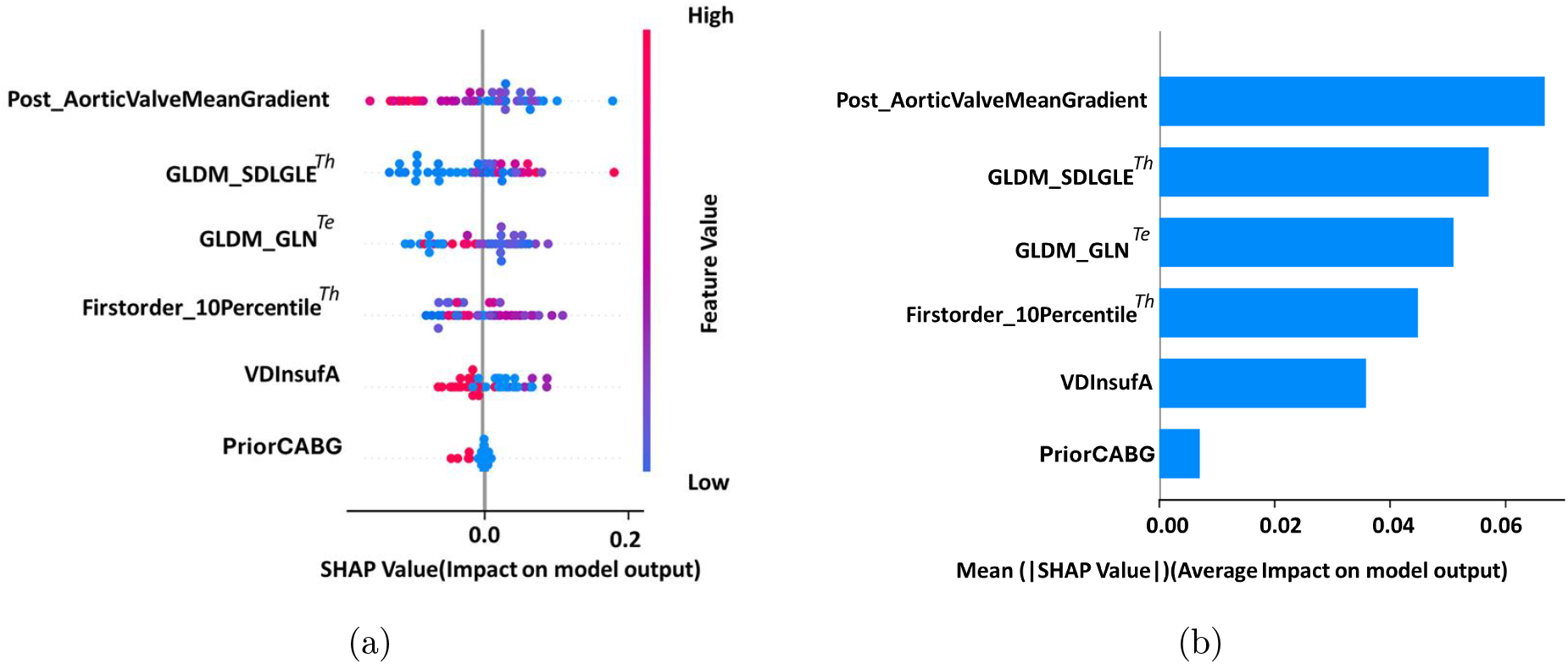
SHAP-Based Feature Importance Analysis. (a) Beeswarm plot showing SHAP values for the top 6 model predictors; each point represents an individual patient, with color indicating normalized feature magnitude (red: high; blue: low) and horizontal position reflecting direction and magnitude of contribution to model output (Adverse remodeling). (b) Mean absolute SHAP values quantifying the average contribution of each feature to model predictions, ranked by importance.

Residual valve insufficiency and prior CABG contributed more modestly. These findings support the complementary role of procedural hemodynamics and imaging-derived markers of regional myocardial heterogeneity in determining post-TAVR remodeling trajectories.

## 4. Discussion

The present study supports the emerging role of routine pre-procedural TAVR CT as a quantitative myocardial phenotyping tool rather than solely an anatomical planning modality. First, CT-derived radiomic markers of myocardial wall-thickness heterogeneity and texture non-uniformity outperformed geometric descriptors alone in identifying patients at risk for adverse LV remodeling after TAVR. Second, a multimodal pre-procedural model integrating CT radiomics with baseline LVMI, valve insufficiency, and revascularization history achieved strong predictive performance (AUC 0.86), supporting the feasibility of pre-procedural risk stratification. Third, incorporation of post-TAVR mean transvalvular gradient further improved discrimination (AUC 0.91), highlighting the incremental prognostic value of post-procedural hemodynamic information and supporting complementary clinical applications of the pre- and post-procedural models. Fourth, SHAP analysis identified residual transvalvular gradient and baseline LVMI as dominant contributors to model predictions, emphasizing the combined influence of hemodynamic burden and myocardial structural remodeling on post-TAVR recovery trajectories.

### 4.1. CT Radiomics and Myocardial Structural Phenotyping

Global LV geometric descriptors, including sphericity and conicity indices, demonstrated limited discriminative performance (AUC 0.62), likely reflecting their inability to capture the regional myocardial heterogeneity associated with maladaptive remodeling under chronic pressure overload. In contrast, radiomic analysis of myocardial wall thickness identified subtle regional thinning patterns and attenuation heterogeneity not captured by conventional geometric measures, resulting in substantially improved discrimination (AUC 0.84). These imaging features may reflect underlying structural remodeling processes, including interstitial fibrosis and cardiomyocyte atrophy, which are known to impair ventricular recovery following valve replacement^14,23^.

The incremental value of myocardial texture and wall-thickness radiomics is consistent with prior studies demonstrating that radiomic signatures can detect subclinical myocardial alterations beyond conventional geometric or echocardiographic parameters. Importantly, the performance achieved in this study is comparable to previously reported CT- and CMR-based radiomics approaches while leveraging routinely acquired pre-procedural CT imaging without additional imaging burden.^24^

### 4.2. Biological Significance of Lower Pre-Procedural LVMI in Adverse Remodeling

The observation that patients with adverse remodeling exhibited significantly lower pre-procedural LVMI (95.39 vs. 116.85 g/m^2^; p<0.0001) may reflect impaired compensatory hypertrophy in response to chronic pressure overload from aortic stenosis(See Figure 5). Lower LVMI could indicate transition toward a more advanced, decompensated myocardial phenotype characterized by fibrosis, cardiomyocyte loss, and reduced contractile reserve, thereby limiting the capacity for reverse remodeling after TAVR. This interpretation is supported by prior CMR studies showing that myocardial fibrosis burden is a stronger predictor of post-TAVR LV mass regression than baseline LVMI alone ^8^. The observed increase in CT texture heterogeneity in adverse remodelers, including higher kurtosis, cluster prominence, and high gray-level run emphasis, further supports the concept that imaging-derived myocardial heterogeneity may reflect biologically relevant structural remodeling processes, although direct histopathologic validation remains necessary.

**Figure 5:**
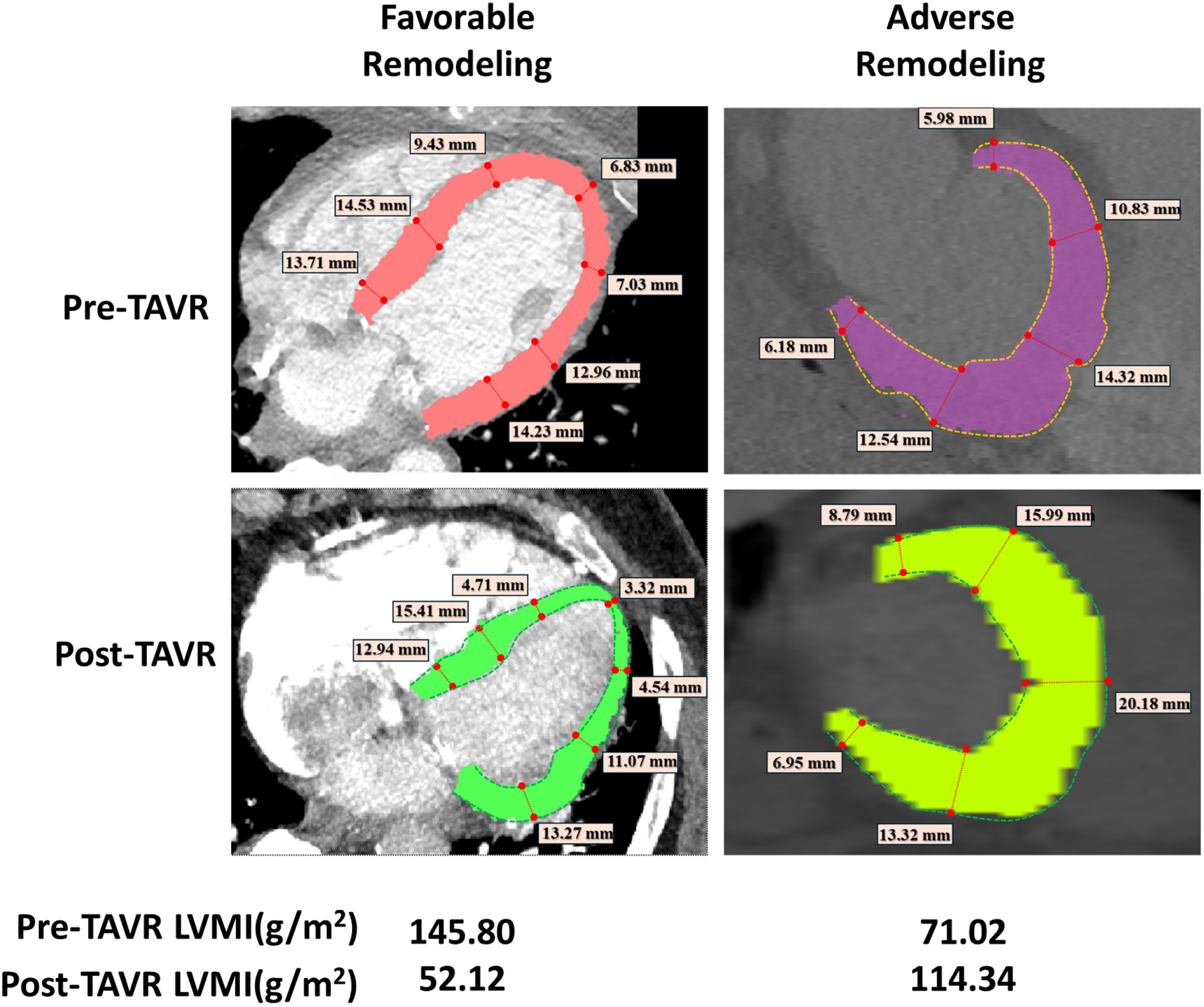
Representative showcase comparison between cases with different post-operative outcomes in terms of LVMI following TAVR. The figure is organized into two columns and two rows, representing favorable remodeling and adverse remodeling cases, as well as pre-TAVR and post-TAVR imaging, respectively. The top row demonstrates pre-TAVR morphology, while the bottom row illustrates post-TAVR follow-up imaging.

### 4.3. The Dominant Role of Residual Haemodynamic Burden

The superior performance of the multimodal model (Model 6; AUC 0.91), which incorporated post-procedural variables, underscores the additive prognostic value of hemodynamic outcomes in determining myocardial recovery after TAVR. Among all predictors, post-procedural mean transvalvular gradient emerged as a key determinant, suggesting that residual afterload substantially influences reverse remodeling. These findings are consistent with prior studies linking elevated residual gradients to adverse remodeling and impaired functional recovery^25^.

The lower pre-procedural LVMI observed in the adverse remodeling cohort may reflect limited hypertrophic reserve and low stroke volume physiology, in which measured gradients can underestimate true residual afterload. When adjusted for baseline LVMI, residual gradient retained independent prognostic significance, supporting its role as a marker of persistent ventricular loading. Persistent post-TAVR gradients, often associated with prosthesis–patient mismatch or incomplete valve expansion, may sustain maladaptive hypertrophic signaling and impair reverse remodeling. These findings highlight the importance of achieving optimal hemodynamic outcomes during TAVR and identifying patients who may benefit from closer post-procedural surveillance or secondary intervention.

### 4.4. Clinical Implications

The proposed two-stage framework may provide clinically actionable risk stratification at distinct stages of TAVR care leveraging imaging data already routinely acquired during standard TAVR evaluation. The pre-procedural model (Model 5), derived from routinely available CT and clinical data, demonstrated high specificity and may support identification of patients at low risk for adverse remodeling, potentially enabling more tailored surveillance strategies. The post-procedural refinement model (Model 6), incorporating residual transvalvular gradient, further improved risk discrimination and may help identify patients who could benefit from closer follow-up or adjunctive therapies targeting adverse remodeling. Although these findings support the potential utility of CT-based radiomic phenotyping for risk stratification, prospective external validation is required before clinical implementation.

Beyond remodeling prediction, radiomic phenotyping may have broader applications in TAVR care. Imaging markers of septal thickness, calcification burden, and myocardial heterogeneity could potentially be integrated with procedural planning tools to identify patients at increased risk for conduction disturbances or pacemaker implantation following TAVR.^26,27^ Similarly, pre-procedural radiomic signatures of hypertrophy and fibrosis may provide incremental information regarding disease staging and timing of intervention. Prior studies from the PARTNER trials demonstrated that failure to achieve LV mass regression after TAVR is associated with substantially higher mortality and re-hospitalization risk. ^7,14^ Radiomic phenotyping may therefore help identify patients with disproportionate structural remodeling who could benefit from earlier intervention, although this hypothesis remains speculative.

Patients identified as higher risk by radiomic profiling may also warrant intensified longitudinal surveillance incorporating advanced echocardiographic indices such as global longitudinal strain or emerging remote monitoring technologies. ^28,29^ In addition, the potential role of radiomic risk phenotyping in guiding treatment selection between TAVR and SAVR remains an area for future investigation, particularly given evidence that SAVR may induce more rapid reverse remodeling in selected patient populations.^30,31^

### 4.5. Threshold Optimization and Clinical Decision Framework

The selected classification thresholds reflect a trade-off between sensitivity and specificity, with higher thresholds favoring specificity and lower thresholds improving sensitivity. The choice of operating threshold should be guided by the intended clinical application, particularly in balancing false positives and false negatives. In this study, thresholds emphasizing high specificity were explored to identify patients at low risk for adverse remodeling with a low false-positive rate, potentially supporting less intensive post-procedural surveillance. However, at the Youden-optimal threshold, Model 5 achieves sensitivity 0.65 and specificity 0.88 (NPV 0.94, PPV 0.52), and Model 6 achieves sensitivity 0.60 and specificity 0.90 (NPV 0.93, PPV 0.55), indicating improved balance between sensitivity and specificity. In clinical practice, the relative consequences of false-negative versus false-positive predictions should be considered, as failure to identify patients at risk for adverse remodeling may be associated with clinically meaningful down-stream events. Approaches to address class imbalance, including resampling techniques, may improve sensitivity and warrant further investigation in future studies.

### 4.6. Study Limitations

Several limitations warrant consideration. The retrospective single-center design and moderate cohort size (n = 232) may limit generalizability, requiring external validation in larger and more diverse multicenter populations. In addition, adverse remodeling occurred in only 22.4% of patients, resulting in class imbalance that may have contributed to reduced sensitivity. Future studies should address this limitation through prospective study design and balanced sampling strategies. Finally, LV remodeling is a dynamic process, and binary classification based on a single follow-up interval may not fully capture longitudinal remodeling trajectories. Future work incorporating serial imaging and continuous remodeling metrics may provide a more comprehensive assessment of ventricular adaptation.

## 5. Conclusions

In this study, CT-derived radiomic markers of myocardial wall-thickness heterogeneity and texture provided greater discrimination of adverse LV remodeling after TAVR than conventional geometric descriptors, highlighting the importance of regional myocardial heterogeneity beyond global chamber morphology. Integration of radiomic features with clinical and post-procedural hemodynamic variables further improved predictive performance, supporting a multifactorial model in which myocardial substrate and residual loading conditions jointly influence remodeling trajectories.

Patients with adverse remodeling demonstrated lower baseline LV mass index and increased radiomic heterogeneity, suggesting that impaired compensatory hypertrophy and underlying myocardial remodeling may limit reverse remodeling after TAVR. The proposed two-stage framework, incorporating both pre- and post-procedural assessment, demonstrates the potential of routinely acquired CT imaging for imaging-informed risk stratification. These findings extend the role of pre-procedural CT beyond anatomical planning toward quantitative myocardial phenotyping, although prospective multicenter validation remains necessary.

### Perspectives

#### Competency In Medical Knowledge

Pre-procedural cardiac CT, routinely acquired for TAVR planning, contains quantitative myocardial information beyond anatomical assessment. CT-derived radiomic characterization of wall-thickness heterogeneity and myocardial texture, integrated with baseline LV mass index and post-procedural transvalvular gradient, identifies patients at risk for adverse LV remodeling and should be considered in post-TAVR risk stratification and surveillance planning.

#### Translational Outlook

A key component of post-TAVR management consists of identifying patients at risk for maladaptive LV remodeling and impaired functional recovery. The present study demonstrates that CT-derived radiomic signatures of myocardial heterogeneity provide incremental prognostic value beyond conventional geometric assessment. Prospective multicenter validation and standardization of radiomic workflows are required, and future studies integrating these imaging biomarkers into existing CT planning platforms could enable seamless, zero-burden risk stratification to guide individualized surveillance and treatment.

## Data Availability

All data produced in the present study are available upon reasonable request to the authors

## Abbreviations

AS: Aortic stenosis
AUC: Area under the curve
CCT: Cardiac computed tomography
CT: Computed tomography
LV: Left ventricular
LVEF: Left ventricular ejection fraction
LVMI: Left ventricular mass index
ML: Machine learning
SHAP: SHapley Additive exPlanations
TAVR: Transcatheter aortic valve replacement

## Data Availability

Imaging data were acquired at Brigham and Women’s hospital (Boston, USA). Derived data supporting this study could be made available by the corresponding author (MR) upon request following the institutional policy.

