## Supplemental Material for "Early Prediction of Post-TAVR Left Ventricular Remodeling Using CT-Derived Radiomics and Clinical Variables"

### S1. Handling of Blank PHI Data

Considering the dataset acquired from Section 2.1, we focused on clinically relevant features previously reported to correlate with post-TAVR LV remodeling outcomes. These features encompassed patient demographics, baseline clinical characteristics, pre-procedural echocardiographic parameters, and post-procedural valve assessment metrics.

Missing data analysis revealed varying degrees of incompleteness across the selected features. To address missing data while optimizing imputation accuracy, we employed an enhanced multivariate imputation strategy. For continuous variables, we implemented KNN imputation with $k = 5$ neighbors across the entire 932-patient dataset[1]. Following imputation across the full 932-patient dataset, we extracted the imputed values specifically for our 232-patient BWH study cohort for all subsequent analyses.

To validate this enhanced imputation approach, we performed cross-validation testing specifically on our 232 BWH patients. We artificially removed 20% of observed values from the cohort, performed imputation using the full 932-patient dataset, and compared imputed values against true values using standard metrics: Root Mean Squared Error (RMSE), Mean Absolute Error (MAE), and Mean Absolute Percentage Error (MAPE)[1].

### S2. Automated Segmentation of the LV Myocardium

LV myocardial segmentation was performed using CardioVision, an in-house platform built on the nnU-Net (v2) framework[2,3]. All contrast-enhanced CT images were preprocessed with intensity normalization and resampling to a standardized spatial resolution to ensure consistent representation across the study cohort. A 3D full-resolution nnU-Net model was trained using 5-fold cross-validation, and final segmentations were obtained by ensemble inference across folds, followed by standard nnU-Net postprocessing to refine masks and remove small spurious regions.

Representative examples are shown in Figure S1. Based on the previously validated segmentation accuracy and low interobserver variability of this AI-based approach[4], the LV myocardial masks were used for downstream geometric measurements, wall-thickness mapping, and radiomic feature extraction. Additional technical details are provided in the Supplementary Materials.

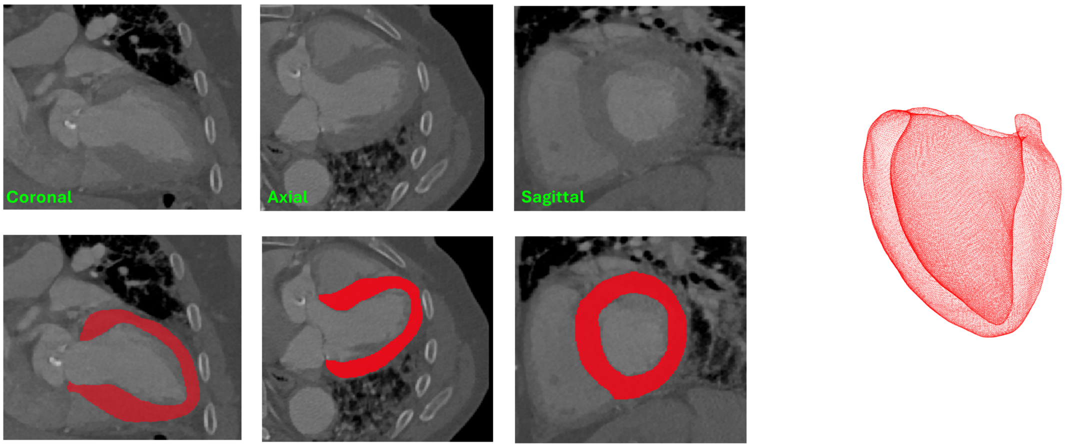


Figure S1: Representative ROI segmentation of the myocardial wall. Left panels show coronal, axial, and sagittal CT slices (top row) and their corresponding segmented masks (bottom row). The right panel depicts the resulting 3D reconstruction of the myocardial geometry.

### S3. Feature Selection and Dimensionality Reduction

Geometric descriptors, myocardial thickness indices, tissue-level radiomic features, and clinical variables were combined to form a high-dimensional feature set. To mitigate overfitting and improve model generalizability, a structured feature selection and dimensionality reduction workflow was implemented in Python.

Candidate variables were first screened using univariate statistical analysis. Continuous variables were compared using the Mann–Whitney U test[5], and categorical variables using the chi-square test[6]. Features with minimal univariate association with adverse LV remodeling ($p > 0.8$) were excluded to reduce noise, while clinically relevant variables were retained irrespective of statistical significance. Permutation-based feature importance was subsequently used to further refine the feature set and reduce redundancy.
